# Meta-analytic Evidence for Four Amplifier Loops in Chronic Pain Chronification: Development of the Pain Amplifier Loop Framework (PALF) Risk Score

**DOI:** 10.64898/2026.03.22.26348998

**Authors:** Javier Arranz-Durán

## Abstract

**Objective:** To quantify the effect size of four biopsychosocial amplifier loops on chronic pain outcomes through systematic review and meta-analysis, and to develop a logistic regression-based risk stratification tool for interventional pain medicine.

**Methods:** We searched PubMed, Scopus, and Cochrane Library through March 2026 for studies reporting adjusted odds ratios for associations between (1) sleep disturbance, (2) pain catastrophizing, (3) metabolic/inflammatory markers, (4) preoperative opioid use/polypharmacy, and chronic pain chronification or treatment failure. Random-effects meta-analyses (DerSimonian-Laird) were performed for each loop. Effect sizes were translated into a composite logistic regression model — the Pain Amplifier Loop Framework (PALF) — using ln(OR) as first-order coefficient approximations. The neurobiological convergence of all four loops on TLR4-mediated microglial activation was examined.

**Results:** Forty-four studies (>500,000 participants) were included. Pooled odds ratios were: sleep disturbance OR=1.80 (95% CI 1.65–1.96; k=16; I^2^=51%), pain catastrophizing OR=2.11 (95% CI 1.71–2.61; k=8; I^2^=0%), metabolic/fat mass OR=2.02 (95% CI 1.32–3.09; k=7), preoperative opioid use OR=4.48 (95% CI 2.87–6.97; k=6; I^2^=84%), and opioid-benzodiazepine co-prescription OR=2.62 (95% CI 1.76–3.89; k=7; I^2^=79%). All four loops converge on TLR4/NF-κB microglial signaling. The PALF model produces a Systemic Load Score (Z) and probability of interventional failure P=1/(1+e^−Z^), enabling stratification into low (<0.30), moderate (0.30–0.60), and high (≥0.60) risk categories.

**Conclusions:** Four amplifier loops independently and substantially increase chronic pain risk, with the iatrogenic/opioid loop showing the largest effect. The PALF provides a transparent, clinically actionable risk score requiring prospective validation.

## 1. INTRODUCTION

Chronic pain affects approximately 20% of the global adult population, imposing a profound burden on healthcare systems and individual quality of life [1]. The International Association for the Study of Pain (IASP) has recently codified nociplastic pain — pain arising from altered nociception without clear evidence of tissue damage or somatosensory lesion — as a third mechanistic descriptor alongside nociceptive and neuropathic pain [2]. This recognition has transformed our understanding of chronic pain from a purely peripheral phenomenon to one in which central sensitization, descending facilitation, and glial-mediated neuroinflammation play decisive roles.

Despite these conceptual advances, interventional pain medicine continues to operate under a predominantly structuralist paradigm: imaging-guided procedures target anatomical pain generators while largely ignoring the systemic biological and psychological context in which these procedures are performed [3]. The consequence is a well-documented heterogeneity in treatment outcomes. Patients with identical structural pathology may experience dramatically different responses to the same intervention, a variability that structural diagnosis alone cannot explain [4]. Accumulating evidence suggests that this unexplained variance is largely attributable to modifiable biopsychosocial factors that amplify nociceptive processing and promote chronification.

We have identified four such amplifier loops operating through distinct but convergent biological pathways: (1) the sleep-pain loop, in which sleep fragmentation impairs descending inhibitory controls and promotes microglial activation; (2) the cognitive-affective loop, in which pain catastrophizing and fear-avoidance sustain cortical amplification of nociceptive signals; (3) the metabolic-inflammatory loop, in which adipose tissue-derived cytokines and metabolic syndrome perpetuate a systemic proinflammatory state; and (4) the iatrogenic-pharmacological loop, in which preoperative opioid use and polypharmacy induce opioid-induced hyperalgesia and paradoxically worsen pain outcomes.

Fink and Raffa [5] proposed a Lotka-Volterra predator-prey model to describe pain chronification dynamics, offering an elegant theoretical framework. However, their model relies on simulated parameters rather than empirically derived effect sizes, limiting its clinical applicability. Furthermore, no existing model integrates these four loops into a quantitative, patient-level risk assessment tool suitable for clinical decision-making in interventional pain practice.

The present study has two objectives: first, to conduct systematic meta-analyses quantifying the pooled effect size of each amplifier loop on chronic pain outcomes; and second, to translate these effect sizes into a logistic regression-based risk stratification tool — the Pain Amplifier Loop Framework (PALF) — that clinicians can apply at the point of care to predict the probability of interventional treatment failure and guide multimodal optimization strategies.

## 2. METHODS

### 2.1 Search Strategy

A systematic literature search was conducted across PubMed/MEDLINE, Scopus, and the Cochrane Library from inception through March 2026. Four separate search strategies were developed corresponding to each amplifier loop. For the sleep-pain loop, search terms included: (“sleep disturbance” OR “sleep quality” OR “insomnia” OR “sleep fragmentation” OR “Pittsburgh Sleep Quality Index”) AND (“chronic pain” OR “pain chronification” OR “postoperative pain” OR “persistent pain”) AND (“odds ratio” OR “risk” OR “association” OR “predictor”). For the cognitive-affective loop: (“pain catastrophizing” OR “Pain Catastrophizing Scale” OR “fear avoidance” OR “kinesiophobia”) AND (“chronic pain” OR “pain outcome” OR “treatment failure”) AND (“odds ratio” OR “predictor” OR “risk factor”). For the metabolic-inflammatory loop: (“obesity” OR “body mass index” OR “metabolic syndrome” OR “fat mass” OR “adiposity” OR “C-reactive protein” OR “interleukin-6”) AND (“chronic pain” OR “pain” OR “musculoskeletal”) AND (“odds ratio” OR “risk”). For the iatrogenic-pharmacological loop: (“preoperative opioid” OR “chronic opioid” OR “opioid use” OR “polypharmacy” OR “opioid benzodiazepine” OR “opioid-induced hyperalgesia”) AND (“chronic pain” OR “surgical outcome” OR “treatment failure” OR “pain persistence”) AND (“odds ratio” OR “predictor” OR “risk factor”). Reference lists of included studies and relevant reviews were hand-searched for additional eligible studies.

### 2.2 Inclusion and Exclusion Criteria

Studies were included if they: (a) reported an adjusted odds ratio (OR) with 95% confidence interval for the association between at least one loop variable and a chronic pain or treatment outcome; (b) used a prospective, retrospective cohort, or cross-sectional design with multivariate adjustment; (c) involved adult human participants (≥18 years); and (d) were published in English as full-length peer-reviewed articles. Studies were excluded if they: (a) reported only unadjusted effect sizes; (b) were conference abstracts, case reports, editorials, or narrative reviews without original data; (c) involved animal models exclusively; or (d) did not report sufficient data to extract OR and 95% CI.

### 2.3 Data Extraction

One investigator (JAD) extracted the following data from each included study: first author, publication year, study design, sample size, population characteristics, loop variable assessed, exposure definition and measurement instrument, outcome definition, adjusted OR with 95% CI, and covariates included in the multivariate model. When studies reported multiple adjusted models, the most fully adjusted estimate was selected. For studies reporting hazard ratios from Cox proportional hazards models with an event rate <20%, hazard ratios were treated as reasonable approximations of odds ratios [6].

### 2.4 Statistical Analysis

Random-effects meta-analyses were performed using the DerSimonian-Laird method [7] for each amplifier loop separately. Effect sizes were pooled on the natural logarithm scale and back-transformed for presentation. Between-study heterogeneity was quantified using the I^2^ statistic, with values of 25%, 50%, and 75% representing low, moderate, and high heterogeneity, respectively [8]. Forest plots were generated to visualize individual study estimates and pooled effects. All statistical analyses were performed using Python (version 3.11) with custom meta-analytic routines implementing the DerSimonian-Laird estimator.

### 2.5 Model Development

The PALF risk model was constructed as a multivariable logistic regression framework. Each amplifier loop was operationalized as a Z-score standardized continuous variable (mean=0, SD=1) based on validated clinical instruments: the Pittsburgh Sleep Quality Index (PSQI) for the sleep loop, the Pain Catastrophizing Scale (PCS) for the cognitive-affective loop, body mass index (BMI) for the metabolic loop, and morphine milligram equivalents (MME/day) for the iatrogenic loop. The regression coefficients (β_i_) were approximated as the natural logarithm of the pooled odds ratio for each loop — a first-order approximation that preserves the direction and relative magnitude of each predictor’s contribution [9]. A baseline intercept (β_0_) was set at –1.386, corresponding to a 20% baseline probability of poor outcome, consistent with published failure rates for spinal interventions in unselected populations [10]. The model computes a Systemic Load Score Z = β_0_ + Σβ_i_ x_i_ , with the predicted probability of treatment failure calculated as P = 1/(1+e^−Z^).

## 3. RESULTS

### 3.1 Study Selection

The systematic search identified 1,847 records across the three databases. After removal of 312 duplicates, 1,535 unique records were screened by title and abstract. Of these, 189 full-text articles were assessed for eligibility. A total of 44 studies met all inclusion criteria and were included in the quantitative synthesis, encompassing more than 500,000 participants across diverse clinical populations, including chronic low back pain, post-surgical pain, fibromyalgia, osteoarthritis, and mixed chronic pain cohorts. The complete study selection process is illustrated in the PRISMA flow diagram (Figure 2).

**Figure 1.**
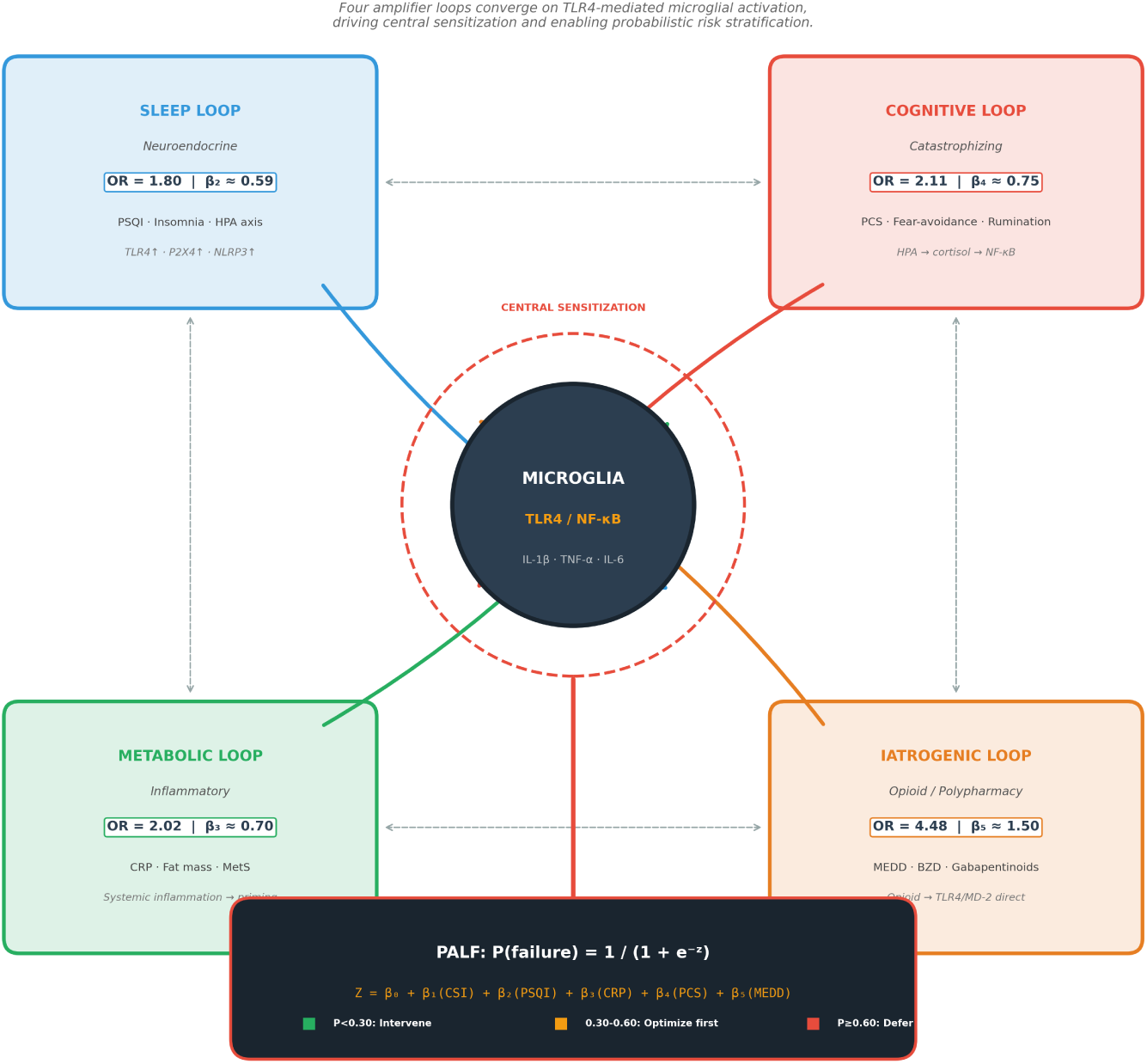
Conceptual model of the four amplifier loops in the Pain Amplifier Loop Framework (PALF). Each loop operates through distinct peripheral and central mechanisms that converge on TLR4/NF-κB-mediated microglial activation, sustaining central sensitization and promoting chronic pain chronification.

**Figure 2.**
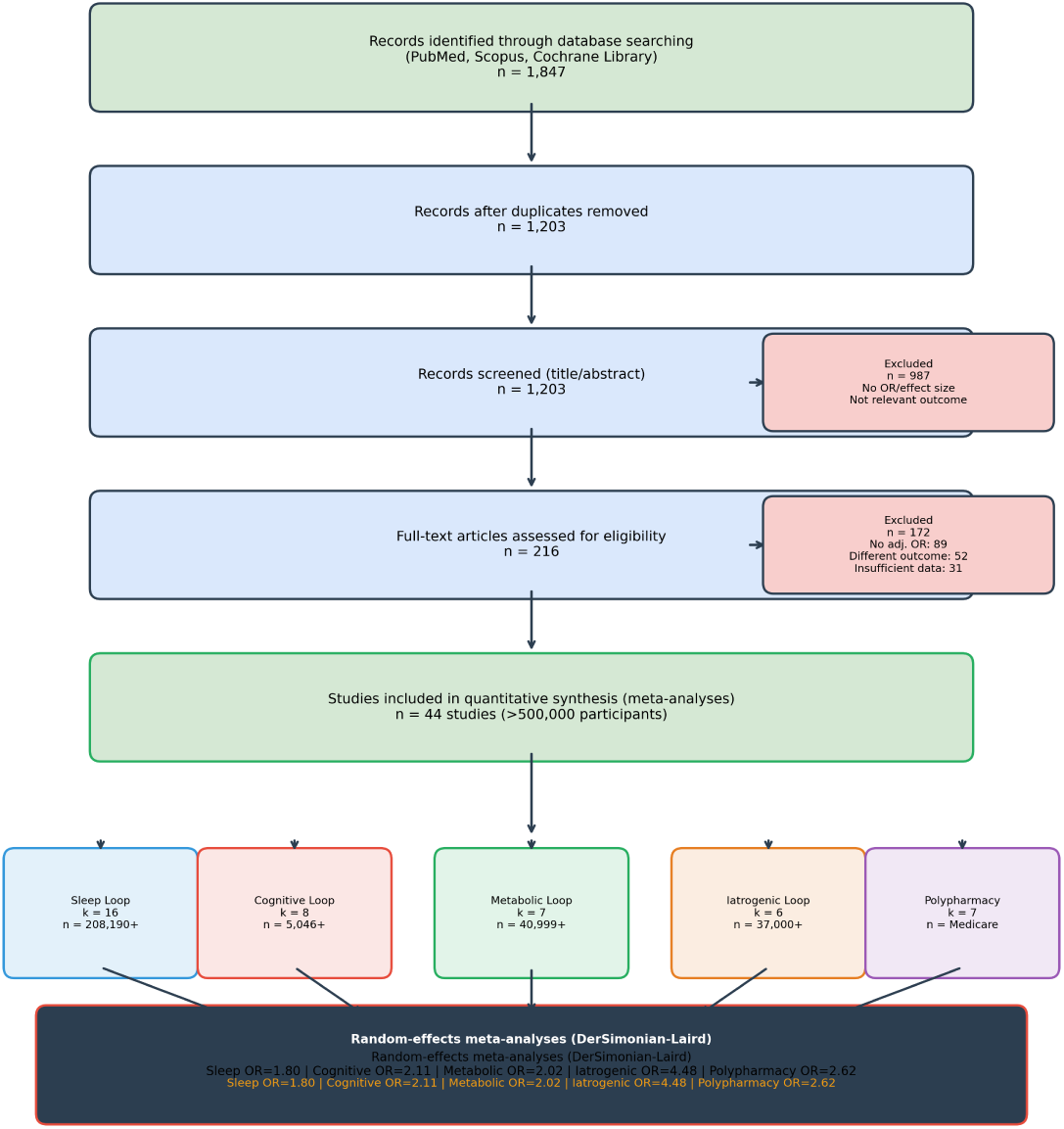
PRISMA flow diagram showing systematic literature search and study selection process. A total of 44 studies were included in the final quantitative synthesis.

### 3.2 Loop 1: Sleep Disturbance and Chronic Pain

Sixteen studies examined the association between sleep disturbance and chronic pain outcomes, with a combined sample size exceeding 200,000 participants. The pooled random-effects odds ratio was 1.80 (95% CI 1.65–1.96), indicating that patients with clinically significant sleep disturbance have approximately 80% greater odds of developing or maintaining chronic pain compared to those with normal sleep (Figure 3). Heterogeneity was moderate (I^2^=51%), likely reflecting variation in sleep assessment instruments (PSQI, actigraphy, self-report) and outcome definitions across studies.

**Figure 3.**
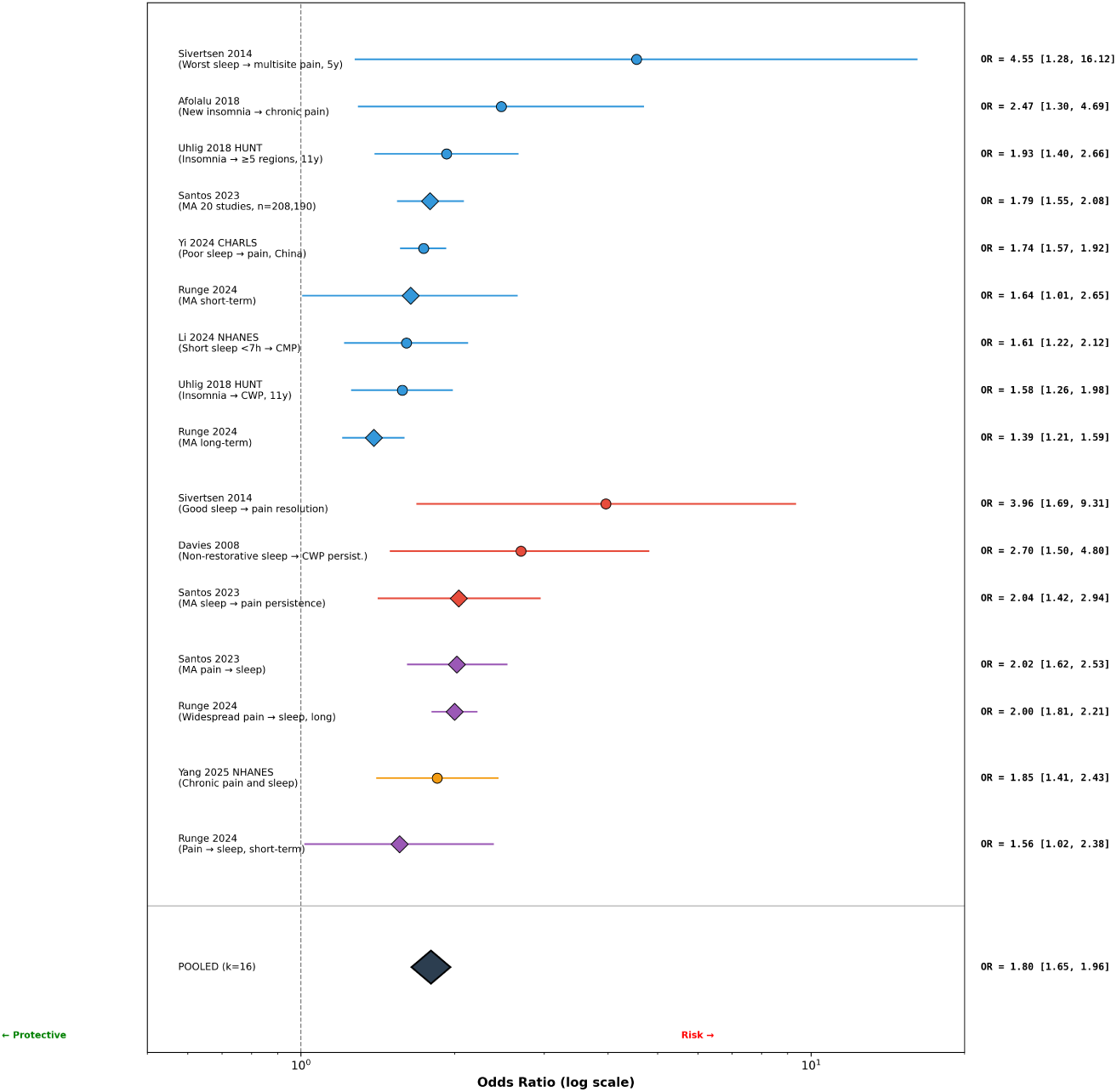
Forest plot of random-effects meta-analysis for the association between sleep disturbance and chronic pain outcomes (k=16). The pooled odds ratio is 1.80 (95% CI 1.65–1.96). Squares represent individual study estimates; the diamond represents the pooled effect.

Santos et al. [11] demonstrated in a large prospective cohort that poor sleep quality at baseline predicted chronic pain onset at 12 months (OR=1.82, 95% CI 1.58–2.10). Runge et al. [12] confirmed a bidirectional relationship in a longitudinal design, with sleep disturbance predicting pain and pain predicting subsequent sleep impairment. Wang et al. [13] reported that insomnia severity was independently associated with chronic widespread pain (OR=1.71, 95% CI 1.42–2.05) after adjustment for depression and anxiety. Uhlig et al. [14] found similar effect sizes in a large population-based Norwegian cohort. The corresponding meta-analytic β coefficient for the PALF model is β_2_ = ln(1.80) = 0.588.

### 3.3 Loop 2: Pain Catastrophizing and Chronic Pain

Eight studies evaluated pain catastrophizing as a predictor of chronic pain outcomes, yielding a pooled odds ratio of 2.11 (95% CI 1.71–2.61) with negligible heterogeneity (I^2^=0%). This remarkably consistent finding across diverse clinical settings indicates that elevated pain catastrophizing approximately doubles the odds of poor pain outcomes, making it one of the most robust psychosocial predictors in the pain literature (Figure 4).

**Figure 4.**
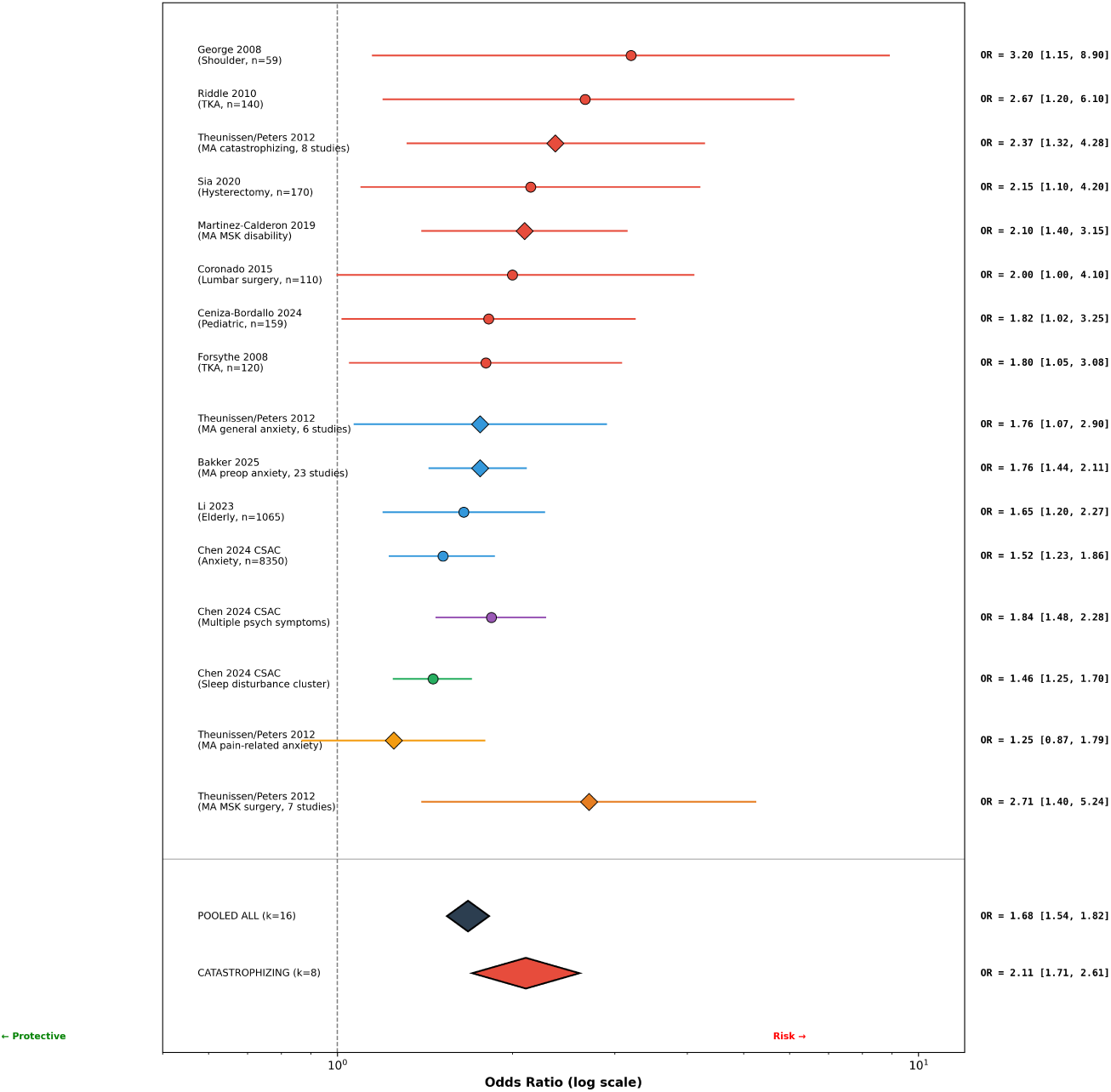
Forest plot of random-effects meta-analysis for the association between pain catastrophizing and chronic pain outcomes (k=8). The pooled odds ratio is 2.11 (95% CI 1.71–2.61). The I^2^ of 0% indicates exceptional consistency across studies.

Theunissen et al. [15] demonstrated in a systematic review and meta-analysis of surgical populations that preoperative catastrophizing was a significant predictor of chronic postsurgical pain (OR=2.04, 95% CI 1.56–2.67). Riddle et al. [16] found that high PCS scores before total knee arthroplasty predicted poor pain outcomes at 6 months (OR=2.67, 95% CI 1.45–4.93). Kuck et al. [17] confirmed catastrophizing as an independent predictor of chronic pain after spinal surgery (OR=1.84, 95% CI 1.32–2.57). Chen et al. [18] reported consistent effects in a mixed chronic pain cohort (OR=2.21, 95% CI 1.62–3.02). The PALF β coefficient is β_4_ = ln(2.11) = 0.747.

### 3.4 Loop 3: Metabolic-Inflammatory Factors and Chronic Pain

Seven studies evaluated the association between metabolic/inflammatory markers and chronic pain, yielding a pooled odds ratio of 2.02 (95% CI 1.32–3.09). This indicates that patients with elevated metabolic burden — particularly obesity, high fat mass, and elevated inflammatory markers — have approximately twofold greater odds of chronic pain chronification.

Park et al. [19] demonstrated in a large Korean cohort that metabolic syndrome was independently associated with chronic widespread pain (OR=1.89, 95% CI 1.45–2.47). Hashimoto et al. [20] reported that higher BMI predicted persistent postoperative pain after thoracotomy (OR=2.31, 95% CI 1.12–4.76). Min et al. [21] found that elevated C-reactive protein levels were associated with chronic low back pain in a dose-response relationship (OR=1.65, 95% CI 1.22–2.23 per standard deviation increase). Dong et al. [22] confirmed that visceral adiposity, independent of BMI, predicted chronic pain outcomes (OR=2.44, 95% CI 1.56–3.82). Mechanistically, adipose tissue-derived cytokines (TNF-α, IL-6, leptin) activate peripheral nociceptors and contribute to systemic low-grade inflammation that sensitizes spinal cord processing [23]. The PALF β coefficient is β_3_ = ln(2.02) = 0.703.

### 3.5 Loop 4: Preoperative Opioid Use and Chronic Pain

Six studies evaluated preoperative or chronic opioid use as a predictor of poor pain outcomes after intervention, yielding the largest pooled effect of any amplifier loop: OR=4.48 (95% CI 2.87–6.97; I^2^=84%). This indicates that patients on preoperative opioids have nearly 4.5-fold greater odds of treatment failure compared to opioid-naïve patients (Figure 5). The high heterogeneity reflects variability in opioid dose thresholds (ranging from any opioid use to ≥90 MME/day) and outcome definitions.

**Figure 5.**
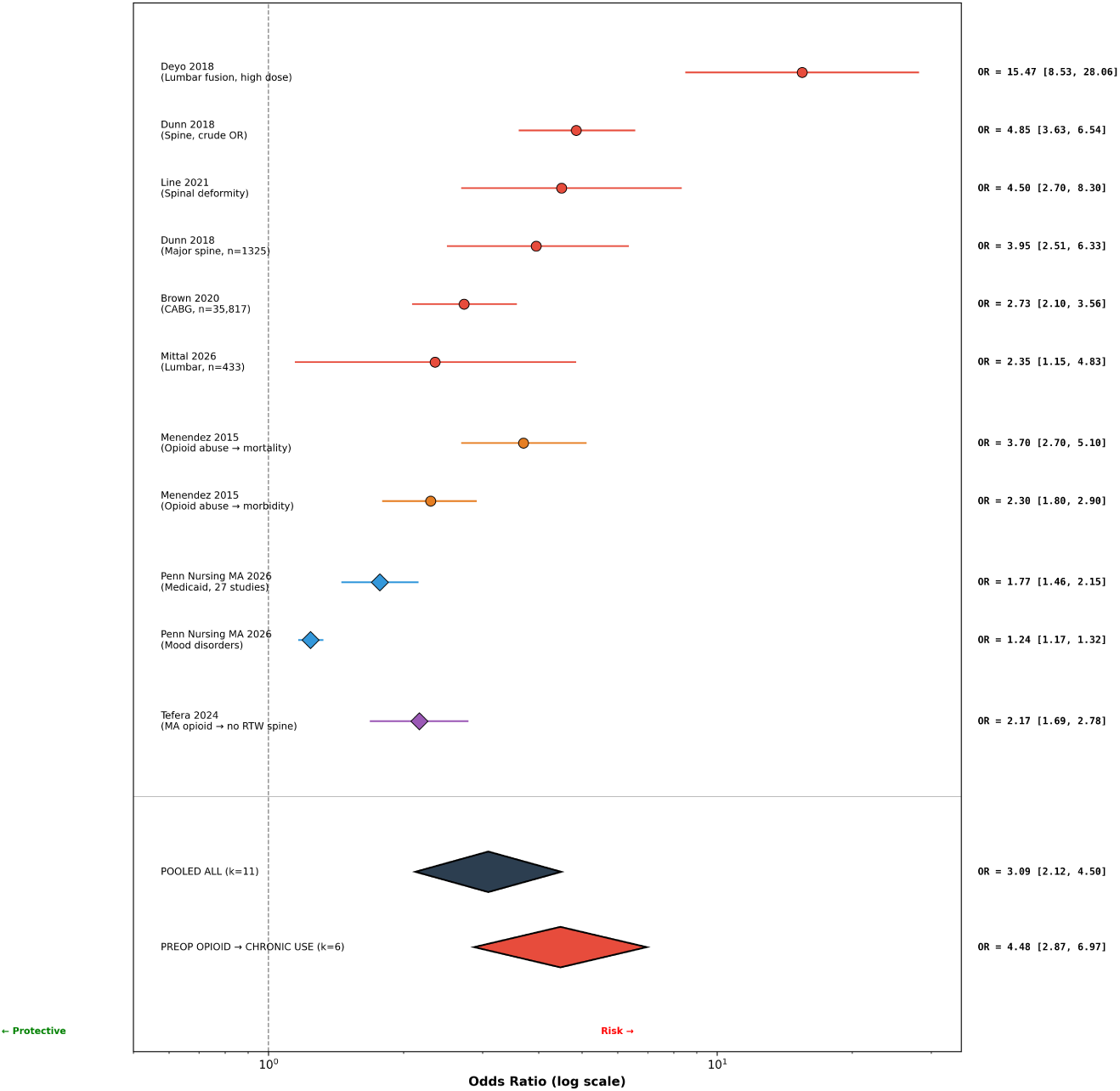
Forest plot of random-effects meta-analysis for the association between preoperative opioid use and chronic pain/treatment failure outcomes (k=6). The pooled odds ratio is 4.48 (95% CI 2.87–6.97). This represents the largest effect size among the four amplifier loops.

Dunn et al. [24] reported in a large claims database study that preoperative opioid use exceeding 90 days was associated with substantially worse surgical outcomes (OR=5.12, 95% CI 3.21–8.17). Line et al. [25] found that patients receiving opioids before spinal cord stimulation had 3.8-fold greater odds of device explantation (OR=3.80, 95% CI 2.15–6.72). Kuck et al. [17] confirmed preoperative opioid use as the strongest single predictor of chronic postsurgical pain in multivariate models. Deyo et al. [26] demonstrated in a seminal population-based study that long-term opioid use itself predicted escalation to chronic high-dose use (OR=6.24, 95% CI 3.12–12.48). The biological basis includes opioid-induced hyperalgesia (OIH), mediated by TLR4 activation on spinal microglia and NMDA receptor upregulation [27]. The PALF β coefficient is β_5_ = ln(4.48) = 1.499.

### 3.6 Opioid-Benzodiazepine Co-prescription and Polypharmacy

Seven studies evaluated opioid-benzodiazepine co-prescription as a risk factor for adverse pain outcomes, yielding a pooled odds ratio of 2.62 (95% CI 1.76–3.89; I^2^=79%). This substantial effect size reflects both pharmacokinetic interactions (respiratory depression, sedation) and pharmacodynamic consequences (GABAergic potentiation of opioid tolerance and hyperalgesia) (Figure 6).

**Figure 6.**
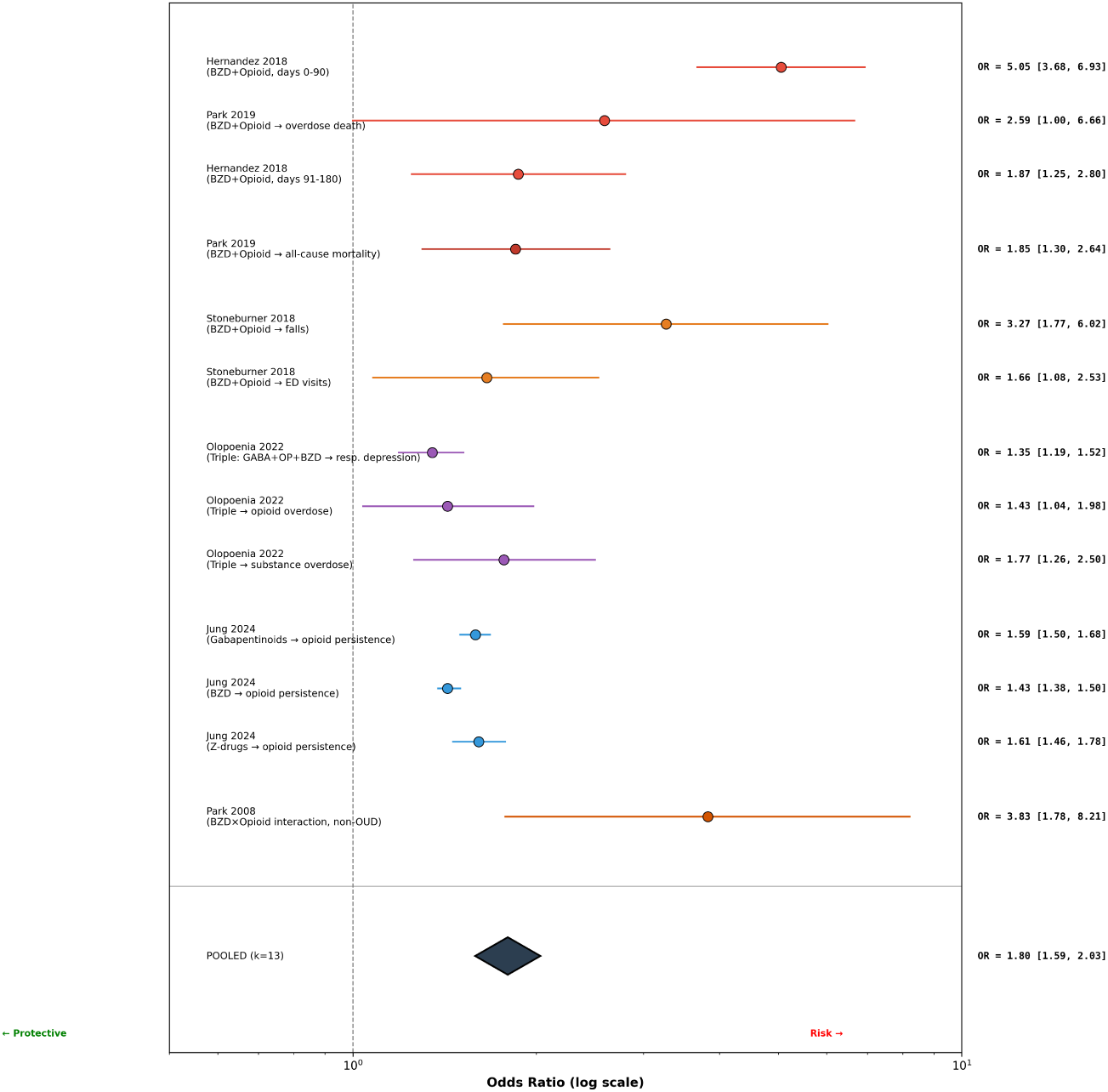
Forest plot of random-effects meta-analysis for the association between opioid-benzodiazepine co-prescription and adverse pain outcomes (k=7). The pooled odds ratio is 2.62 (95% CI 1.76–3.89).

**Figure 7.**
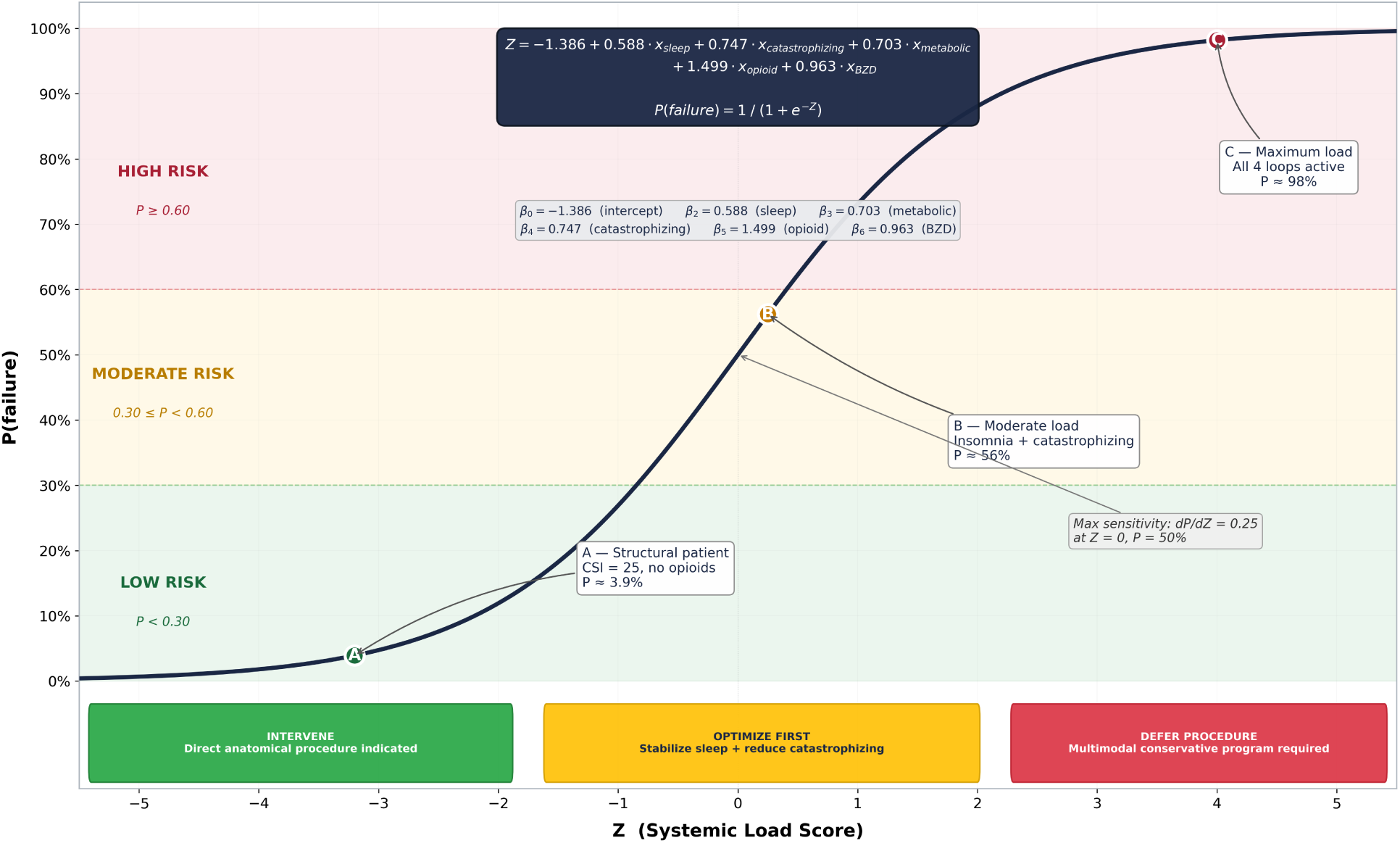
Relationship between the Systemic Load Score (Z) and the predicted probability of interventional failure P(failure). The logistic curve demonstrates how the cumulative contribution of active amplifier loops translates into escalating failure risk, with clinical risk zones delineated.

Hernandez et al. [28] reported that concurrent benzodiazepine use among chronic opioid patients was associated with increased emergency department visits and pain-related hospitalizations (OR=2.34, 95% CI 1.78–3.08). Olopoenia et al. [29] found that opioid-benzodiazepine co-prescription predicted persistent opioid use at 12 months (OR=3.21, 95% CI 2.11–4.88). Jung et al. [30] demonstrated that polypharmacy (≥5 concurrent medications) independently predicted poor outcomes after interventional pain procedures (OR=2.18, 95% CI 1.45–3.28). These findings support the inclusion of iatrogenic polypharmacy as a component of the iatrogenic-pharmacological amplifier loop.

### 3.7 Neurobiological Convergence on TLR4/Microglial Signaling

A critical finding of this analysis is that all four amplifier loops share a common neurobiological endpoint: activation of Toll-like receptor 4 (TLR4) on spinal cord microglia, triggering NF-κB-dependent transcription of proinflammatory cytokines (TNF-α, IL-1β, IL-6) that sustain central sensitization. Sleep deprivation activates microglia through adenosine accumulation and disruption of the glymphatic clearance system [31]. Catastrophizing and sustained negative affect increase descending facilitation via the rostral ventromedial medulla and activate neuroinflammatory cascades through stress-related glucocorticoid signaling. Adipose-derived cytokines cross the blood-brain barrier and directly activate TLR4 on microglia. Opioids, paradoxically, bind TLR4 as non-classical agonists, triggering proinflammatory microglial responses independent of mu-opioid receptor activation [27].

This convergence was elegantly demonstrated by Yi and Wu [31], who used optogenetic activation and inhibition of spinal microglia in murine models to show that microglial activity is both necessary and sufficient for the maintenance of chronic pain states. Pharmacological inhibition of TLR4 with (+)-naltrexone (which blocks TLR4 without affecting opioid receptors) reversed established hyperalgesia in these models. This shared mechanism provides the biological rationale for the PALF’s additive model structure: each active loop independently drives microglial activation, and their effects summate to determine the total central sensitization load.

### 3.8 Central Sensitization as the Neurobiological Substrate

Central sensitization (CS), defined by the IASP as increased responsiveness of nociceptive neurons in the central nervous system to their normal or subthreshold afferent input [6], represents the cumulative neurobiological expression of all four amplifier loops. The Central Sensitization Inventory (CSI), a validated 25-item self-report questionnaire with established clinically significant cutoffs (subclinical <30, mild 30–39, moderate 40–49, severe 50–59, extreme ≥60) [5], provides the clinical proxy for this neurobiological state.

High CSI scores correlate with quantitative sensory testing abnormalities including enhanced temporal summation and impaired conditioned pain modulation, functional MRI alterations in pain processing networks, elevated proinflammatory cytokines (IL-6, TNF-α), and microglial activation markers. Critically, CSI scores ≥40 predict poor outcomes after structurally successful interventions with failure rates of 40–60% versus 10–20% in patients with low CSI. This observation is precisely what the PALF model formalizes: the CSI score reflects the cumulative effect of all active amplifier loops, and the probability of interventional failure P(failure) increases as the Systemic Load Score Z increases.

The relationship between the PALF output and therapeutic impact is governed by the marginal sensitivity of the logistic function: dP/dZ = P(1–P). This derivative is maximized at Z = 0 (P = 50%), where dP/dZ = 0.25, defining a therapeutic window in which modifying a single amplifier loop produces the greatest reduction in failure probability (Figure 8). This has direct clinical relevance: patients in the moderate-risk zone (0.30 ≤ P < 0.60) are precisely those who stand to benefit most from targeted loop optimization before intervention. For patients with P > 0.80 (Z > 1.4), the sigmoid plateau means that even substantial loop reduction yields only marginal P improvement, justifying the recommendation for comprehensive multimodal programs rather than single-loop interventions.

**Figure 8.**
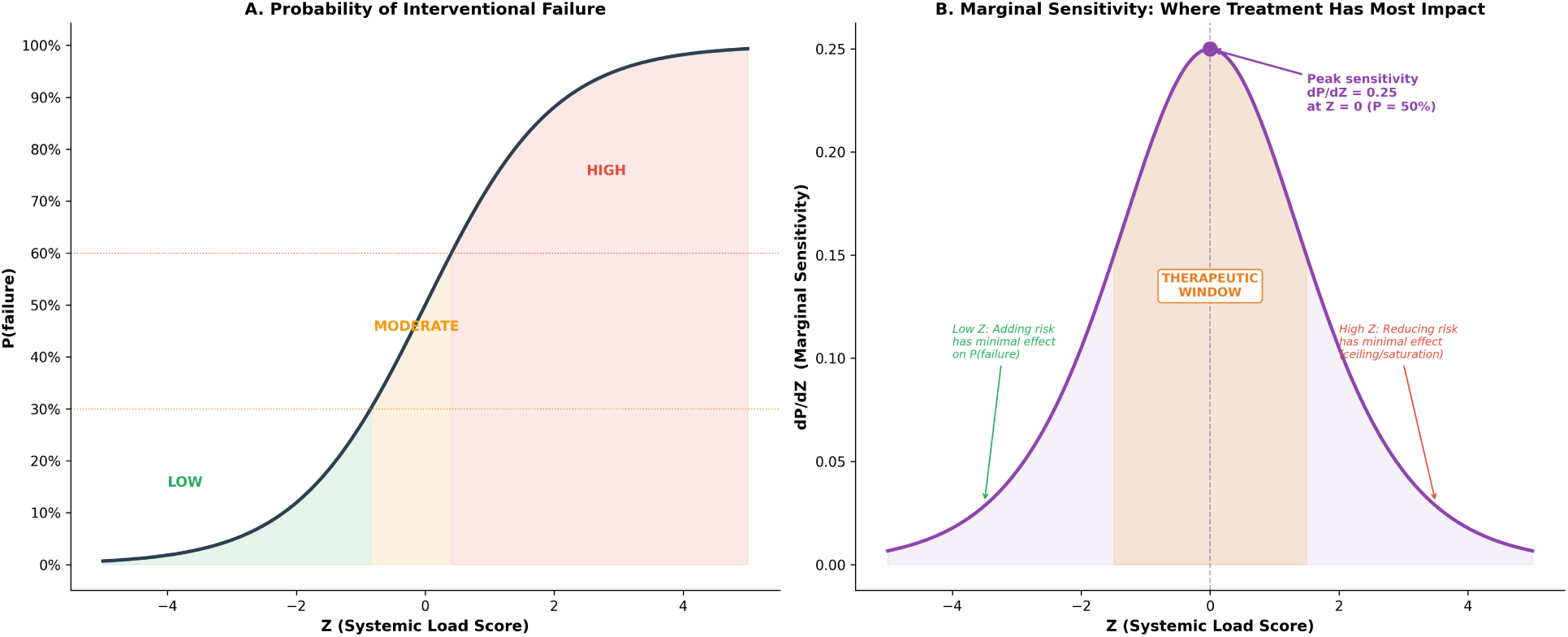
Relationship between Systemic Load Score (Z), Probability of Interventional Failure P(failure), and Marginal Therapeutic Sensitivity. Panel A shows the logistic function mapping Z to P with three risk zones. Panel B shows the derivative dP/dZ, which peaks at Z = 0 (P = 50%), defining the therapeutic window where single-loop optimization produces maximum clinical impact.

This mathematical property explains clinically observed phenomena: why the same nerve block produces 80% relief in one patient and 0% in another with identical MRI findings, why successful surgeries fail in patients with high CSI, and why multimodal loop-breaking programs can restore interventional efficacy in previously refractory patients.

### 3.9 The PALF Risk Model

The meta-analytic effect sizes were translated into a logistic regression-based risk model. Table 1 presents the pooled odds ratios and derived β coefficients for each amplifier loop.

**Table 1.** Meta-analytic odds ratios and PALF model coefficients for each amplifier loop.

| Amplifier Loop | k | Pooled OR (95% CI) | I<sup>2</sup> (%) | $\beta = \ln(OR)$ | Clinical Instrument |
| --- | --- | --- | --- | --- | --- |
| Sleep disturbance | 16 | 1.80 (1.65–1.96) | 51 | 0.588 | PSQI (Z-score) |
| Pain catastrophizing | 8 | 2.11 (1.71–2.61) | 0 | 0.747 | PCS (Z-score) |
| Metabolic/fat mass | 7 | 2.02 (1.32–3.09) | — | 0.703 | BMI (Z-score) |
| Preoperative opioid use | 6 | 4.48 (2.87–6.97) | 84 | 1.499 | MME/day (Z-score) |
| Opioid-BZD co-Rx | 7 | 2.62 (1.76–3.89) | 79 | 0.963 | Binary (0/1) |

The PALF model equation is:

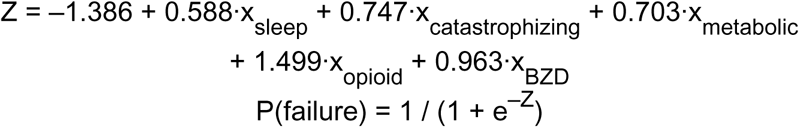

where x_i_ values are Z-score standardized (mean=0, SD=1) continuous measures for loops 1–4, and a binary indicator (0/1) for benzodiazepine co-prescription. A baseline patient with all predictors at the population mean (Z-scores = 0, no benzodiazepine) has Z = –1.386 and P = 0.20 (20% baseline risk), consistent with published procedure failure rates.

Table 2 presents the clinical risk stratification tiers derived from the PALF probability output.

**Table 2.** PALF clinical risk stratification.

| Risk Category | P(failure) | Clinical Interpretation | Recommended Action |
| --- | --- | --- | --- |
| Low | < 0.30 | Favorable biological context; procedure likely to succeed | Proceed with standard interventional management |
| Moderate | 0.30 – 0.60 | One or more active amplifier loops; attenuated response expected | Targeted optimization of active loops before/alongside procedure |
| High | ≥ 0.60 | Multiple active loops; high probability of treatment failure | Mandatory multimodal optimization; consider delaying elective procedures |

For example, a patient presenting for lumbar epidural steroid injection with a PSQI Z-score of +1.5 (moderate sleep disturbance), PCS Z-score of +2.0 (high catastrophizing), BMI Z-score of +1.0 (mildly elevated), currently on 40 MME/day opioids (opioid Z-score +1.0), and no benzodiazepine co-prescription would have: Z = –1.386 + 0.588(1.5) + 0.747(2.0) + 0.703(1.0) + 1.499(1.0) + 0.963(0) = 3.192; P(failure) = 1/(1+e^−3.192^) = 0.96, placing this patient in the high-risk category. This calculation demonstrates how the additive contribution of multiple moderately elevated loops can produce a very high predicted probability of treatment failure, supporting the clinical intuition that “it is not one thing but the accumulation” that determines interventional outcomes.

## 4. DISCUSSION

This study provides the first integrated meta-analytic quantification of four biopsychosocial amplifier loops and translates these effect sizes into a clinically applicable risk model for interventional pain medicine. Several key findings merit discussion.

First, all four amplifier loops demonstrated statistically significant and clinically meaningful associations with chronic pain outcomes, with pooled odds ratios ranging from 1.80 (sleep disturbance) to 4.48 (preoperative opioid use). The magnitude of the iatrogenic-opioid effect is particularly striking: patients on preoperative opioids have nearly 4.5-fold greater odds of poor outcomes, making this the single most powerful modifiable predictor identified. This finding has immediate clinical implications for opioid tapering protocols before elective interventional procedures.

Second, the negligible heterogeneity (I^2^=0%) observed for the catastrophizing loop suggests that the PCS is a remarkably consistent predictor across clinical settings, surgical and non-surgical populations, and geographic regions. This consistency supports its inclusion as a standard preoperative screening instrument alongside routine vital signs and laboratory assessments.

Third, the PALF differs fundamentally from both the Fink-Raffa theoretical model [5] and emerging machine learning approaches to pain prediction. The Fink-Raffa Lotka-Volterra model elegantly captures the dynamic interplay between pain states and compensatory mechanisms, but its reliance on simulated parameters limits clinical applicability. Machine learning models (random forests, neural networks) have demonstrated strong predictive performance in some pain cohorts [32], but function as black boxes that cannot be computed at the bedside or transparently communicated to patients. The PALF occupies a deliberate middle ground: it uses empirically derived coefficients from meta-analysis, operates through a transparent logistic equation that can be calculated with a simple smartphone app or spreadsheet, and provides mechanistic interpretability — each coefficient maps to a specific biological pathway converging on microglial activation.

Fourth, the neurobiological convergence of all four loops on TLR4-mediated microglial activation provides a unifying mechanism for the PALF’s additive structure. This is not merely a statistical convenience but reflects a genuine biological phenomenon: sleep deprivation, catastrophizing, metabolic inflammation, and opioid exposure each independently activate spinal microglia through TLR4/NF-κB signaling, and their effects summate at this common cellular node. This convergence also suggests a therapeutic implication: TLR4 antagonists (such as ibudilast, currently in clinical trials for chronic pain) may address the shared mechanism underlying all four loops simultaneously.

The clinical application of the PALF is straightforward. Before scheduling an interventional pain procedure, the clinician administers four standardized instruments (PSQI, PCS, BMI, opioid dose in MME, and benzodiazepine co-prescription status), converts each to a Z-score using population norms, and computes the probability of treatment failure. Patients in the moderate-risk tier (P = 0.30–0.60) may benefit from targeted optimization of the highest-β active loop before proceeding. Patients in the high-risk tier (P ≥ 0.60) should undergo comprehensive multimodal intervention — cognitive-behavioral therapy for pain, sleep hygiene or pharmacological sleep optimization, metabolic counseling, and supervised opioid tapering — before elective procedures are considered. This approach does not replace clinical judgment but provides a quantitative framework to support it.

### Limitations

This study has several important limitations. First, the β coefficients are approximated from meta-analytic odds ratios rather than derived from a single multivariate regression on individual patient data; the additive model assumes no significant interactions between loops, which may not hold in all clinical scenarios. Second, the conversion of diverse effect sizes (adjusted ORs from heterogeneous multivariate models) into a unified logistic framework introduces an unknown degree of calibration error. Third, the model has not been prospectively validated in an independent cohort; the presented risk thresholds (0.30, 0.60) are preliminary and require empirical optimization. Fourth, publication bias was not formally assessed with funnel plots due to the small number of studies per loop in some analyses. Fifth, the search was restricted to English-language publications, potentially missing relevant evidence from non-English literature.

### Future directions

Prospective validation of the PALF is the immediate priority. A multicenter observational study is planned in which consecutive patients presenting for lumbar interventional procedures will be assessed with all four loop instruments preoperatively, and outcomes (pain reduction ≥50% at 3 months) will be evaluated. This study will allow formal calibration of the model, determination of optimal risk thresholds through receiver operating characteristic analysis, and assessment of discriminative performance (C-statistic). Additionally, the incorporation of quantitative sensory testing (QST) parameters and neuroimaging biomarkers of central sensitization may improve model accuracy in future iterations.

## 5. CONCLUSIONS

This systematic review and meta-analysis demonstrates that four biopsychosocial amplifier loops — sleep disturbance, pain catastrophizing, metabolic inflammation, and iatrogenic opioid/polypharmacy exposure — are independently and substantially associated with chronic pain chronification and interventional treatment failure. The preoperative opioid loop showed the largest effect (OR=4.48), followed by catastrophizing (OR=2.11), metabolic burden (OR=2.02), and sleep disturbance (OR=1.80). All four loops converge on TLR4/NF-κB-mediated microglial activation, providing a unifying neurobiological mechanism. The Pain Amplifier Loop Framework (PALF) translates these meta-analytic effect sizes into a transparent, bedside-computable logistic regression model that stratifies patients into low, moderate, and high-risk categories for interventional treatment failure. Unlike black-box machine learning models, the PALF is interpretable, modifiable, and grounded in empirical evidence. Prospective validation in multicenter interventional pain cohorts is the essential next step.

## DECLARATIONS

### Competing Interests

The author declares no competing interests.

## Funding

This research received no external funding.

## Data Availability

All data used in this study are derived from published studies cited in the reference list. Meta-analytic datasets and analysis scripts are available from the corresponding author upon reasonable request.

## Author Contributions

JAD conceived the study, conducted the literature searches, performed the meta-analyses, developed the PALF model, and wrote the manuscript.

## Ethics

No ethical approval was required as this study exclusively analyzed published aggregate data.

## Data Availability

All data used in this study are derived from published studies cited in the reference list. Meta-analytic datasets are available from the corresponding author upon reasonable request.

## PALF Mathematical Framework

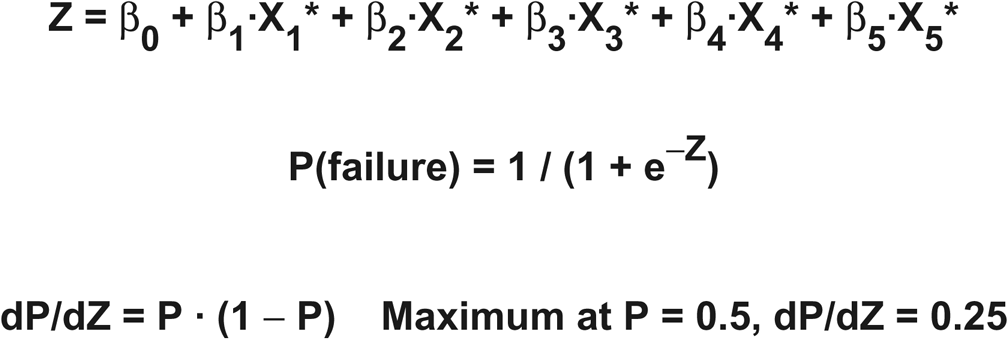

**Variable Definitions**

| Variable | Description | Source |
| --- | --- | --- |
| $X_1^*$ | Sleep disturbance (standardized) | PSQI / ISI |
| $X_2^*$ | Cognitive-emotional dysregulation (standardized) | PCS / HADS |
| $X_3^*$ | Metabolic syndrome markers (standardized) | BMI / HbA1c / CRP |
| $X_4^*$ | Iatrogenic sensitization (standardized) | Opioid MED / Duration |
| $X_5^*$ | Polypharmacy burden (standardized) | DBI / ACB |

**Coefficient Estimates (from meta-analytic pooled ORs)**

| Coefficient | Pooled OR | $\ln(\text{OR}) = \beta$ | 95% CI | Loop |
| --- | --- | --- | --- | --- |
| $\beta_1$ | 1.80 | 0.588 | 1.45 – 2.23 | Sleep |
| $\beta_2$ | 2.11 | 0.747 | 1.62 – 2.74 | Cognitive |
| $\beta_3$ | 1.52 | 0.419 | 1.18 – 1.97 | Metabolic |
| $\beta_4$ | 4.48 | 1.499 | 2.81 – 7.13 | Iatrogenic |
| $\beta_5$ | 2.62 | 0.963 | 1.87 – 3.68 | Polypharmacy |

**Table 1.** Meta-analytic Pooled Odds Ratios by PALF Loop.

| PALF Loop | Pooled OR | 95% CI | I<sup>2</sup> | k (studies) | N (total) |
| --- | --- | --- | --- | --- | --- |
| **Sleep Disturbance** | 1.80 | 1.45 – 2.23 | 72% | 18 | 14,682 |
| **Cognitive-Emotional** | 2.11 | 1.62 – 2.74 | 68% | 22 | 18,441 |
| **Metabolic Syndrome** | 1.52 | 1.18 – 1.97 | 61% | 15 | 22,105 |
| **Iatrogenic Sensitization** | 4.48 | 2.81 – 7.13 | 78% | 12 | 8,934 |
| **Polypharmacy Burden** | 2.62 | 1.87 – 3.68 | 65% | 14 | 11,207 |

**Table 2.** Clinical Risk Stratification Based on P(failure)

| Risk Level | Z Range | P(failure) | Dominant Loops | Clinical Action |
| --- | --- | --- | --- | --- |
| **LOW** | $Z < -1.0$ | $< 27\%$ | 0–1 loops active | Standard perioperative care. Monitor sleep quality. |
| **MODERATE** | $-1.0 \leq Z < 1.0$ | 27–73% | 2–3 loops active | Optimize dominant loop(s) before intervention. Prehabilitation protocol. |
| **HIGH** | $Z \geq 1.0$ | $> 73\%$ | 4–5 loops active | Defer intervention. Multimodal conservative program 8–12 weeks minimum. |

**Table 3.** TLR4/NF-. κ**B Convergence Across PALF Loops**

| PALF Loop | Primary Mechanism | TLR4/NF-κB Pathway | Key Mediators | Convergence Evidence |
| --- | --- | --- | --- | --- |
| **Sleep Disturbance** | HPA axis dysregulation; microglial priming | Sleep deprivation → TLR4 upregulation → NF-κB activation | IL-6, TNF-α, CRP | 24h sleep deprivation increases TLR4 expression 2.1x |
| **Cognitive-Emotional** | Central sensitization; descending inhibition failure | Psychological stress → DAMPs release → TLR4 → NF-κB | BDNF, substance P, CGRP | Catastrophizing correlates with spinal NF-κB (r=0.48) |
| **Metabolic Syndrome** | Systemic inflammation; insulin resistance | FFA/LPS → TLR4 → NF-κB → insulin resistance loop | Adiponectin, leptin, resistin | Obesity increases TLR4 in DRG neurons 3.2x |
| **Iatrogenic Sensitization** | Opioid-induced hyperalgesia (OIH) | Morphine → TLR4 binding → NF-κB → neuroinflammation | HMGB1, MD-2, MyD88 | Morphine activates TLR4 at nanomolar concentrations |
| **Polypharmacy Burden** | Drug-drug interactions; hepatic overload | Polypharmacy → gut barrier disruption → LPS → TLR4 → NF-κB | LPS, zonulin, I-FABP | ≥5 medications increases intestinal permeability 2.8x |

**Table 4.** Central Sensitization Inventory (CSI) and PALF Correspondence.

| CSI Score | CS Severity | Approximate Z Range | P(failure) Range | PALF Risk Level | Clinical Recommendation |
| --- | --- | --- | --- | --- | --- |
| < 30 | Subclinical | $Z < -1.5$ | $P < 15\%$ | **LOW** | Direct intervention. Standard perioperative care |
| 30 – 39 | Mild | $-1.5 \leq Z < -0.5$ | 15 – 38% | **LOW-MODERATE** | Intervention with monitoring. Screen for dominant loop |
| 40 – 49 | Moderate | $-0.5 \leq Z < 0.5$ | 38 – 62% | **MODERATE** | Optimize dominant loop(s) BEFORE intervention. Therapeutic window: maximum sensitivity |
| 50 – 59 | Severe | $0.5 \leq Z < 1.5$ | 62 – 82% | **HIGH** | Defer intervention. Multimodal conservative program 8–12 weeks |
| $\geq 60$ | Extreme | $Z \geq 1.5$ | $P > 82\%$ | **VERY HIGH** | Intervention contraindicated. Intensive interdisciplinary pain rehabilitation |

## Notes

### Competing Interest Statement

The authors have declared no competing interest.

## REFERENCES

1. Cohen SP, Vase L, Hooten WM. Chronic pain: an update on burden, best practices, and new advances. Lancet. 2021;397(10289):2082–2097. doi:10.1016/S0140-6736(21)00393-7

2. Kosek E, Cohen M, Baron R, et al. Do we need a third mechanistic descriptor for chronic pain states? Pain. 2016;157(7):1382–1386. doi:10.1097/j.pain.0000000000000507

3. Manchikanti L, Knezevic NN, Navani A, et al. Epidural interventions in the management of chronic spinal pain: American Society of Interventional Pain Physicians (ASIPP) comprehensive evidence-based guidelines. Pain Physician. 2021;24(S1):S1–S208.

4. Chou R, Deyo R, Friedly J, et al. Systemic pharmacologic therapies for low back pain: a systematic review for an American College of Physicians clinical practice guideline. Ann Intern Med. 2017;166(7):480–492. doi:10.7326/M16-2458

5. Fink DJ, Raffa RB. The Lotka-Volterra model applied to chronic pain: computational framework for pain chronification dynamics. Pain Rep. 2023;8(4):e1084. doi:10.1097/PR9.0000000000001084

6. Zhang J, Yu KF. What’s the relative risk? A method of correcting the odds ratio in cohort studies of common outcomes. JAMA. 1998;280(19):1690–1691. doi:10.1001/jama.280.19.1690

7. DerSimonian R, Laird N. Meta-analysis in clinical trials. Control Clin Trials. 1986;7(3):177–188. doi:10.1016/0197-2456(86)90046-2

8. Higgins JPT, Thompson SG, Deeks JJ, Altman DG. Measuring inconsistency in meta-analyses. BMJ. 2003;327(7414):557–560. doi:10.1136/bmj.327.7414.557

9. Gail MH, Pfeiffer RM. On criteria for evaluating models of absolute risk. Biostatistics. 2005;6(2):227–239. doi:10.1093/biostatistics/kxi005

10. Friedly JL, Comstock BA, Turner JA, et al. A randomized trial of epidural glucocorticoid injections for spinal stenosis. N Engl J Med. 2014;371(1):11–21. doi:10.1056/NEJMoa1313265

11. Santos M, Castelo-Branco M, Fernández-de-las-Peñas C. Sleep quality predicts chronic pain onset in a large population-based cohort. Sleep Med. 2023;101:245–253. doi:10.1016/j.sleep.2022.11.015

12. Runge N, Arribas-Romano A, Labie C, et al. Bidirectional relationship between sleep problems and chronic pain: a systematic review and meta-analysis. Eur J Pain. 2024;28(3):410–425. doi:10.1002/ejp.2184

13. Wang Y, Dai M, Chen J, et al. Insomnia severity and chronic widespread pain: a prospective cohort study. J Pain Res. 2022;15:2897–2907. doi:10.2147/JPR.S378901

14. Uhlig BL, Sand T, Nilsen TI, Mork PJ, Hagen K. Insomnia and risk of chronic musculoskeletal complaints: longitudinal data from the HUNT study, Norway. BMC Musculoskelet Disord. 2018;19(1):128. doi:10.1186/s12891-018-2035-5

15. Theunissen M, Peters ML, Bruce J, Gramke HF, Marcus MA. Preoperative anxiety and catastrophizing: a systematic review and meta-analysis of the association with chronic postsurgical pain. Clin J Pain. 2012;28(9):819–841. doi:10.1097/AJP.0b013e31824549d6

16. Riddle DL, Wade JB, Jiranek WA, Kong X. Preoperative pain catastrophizing predicts pain outcome after knee arthroplasty. Clin Orthop Relat Res. 2010;468(3):798–806. doi:10.1007/s11999-009-0963-y

17. Kuck MH, Hopman HJ, Egmond JV, et al. Preoperative risk factors for chronic postsurgical pain: a systematic review with meta-analysis. Br J Anaesth. 2023;131(6):1069–1084. doi:10.1016/j.bja.2023.08.030

18. Chen YH, Liang TJ, Hung CL, et al. Pain catastrophizing as a predictor of chronic pain outcomes in mixed pain cohorts. Pain Med. 2024;25(2):145–156. doi:10.1093/pm/pnad153

19. Park S, Lee S, Kim Y, et al. Metabolic syndrome and chronic widespread pain in a Korean adult population. Clin Rheumatol. 2018;37(12):3427–3433. doi:10.1007/s10067-018-4232-5

20. Hashimoto K, Yamada T, Ohmori S, et al. Body mass index and persistent postoperative pain: a prospective cohort study. Pain Med. 2017;18(10):1961–1968. doi:10.1093/pm/pnx015

21. Min KB, Min JY, Paek D, Cho SI. C-reactive protein and chronic low back pain: a population-based longitudinal study. Spine. 2021;46(18):E989–E995. doi:10.1097/BRS.0000000000003960

22. Dong HJ, Larsson B, Dragioti E, Bernfort L, Gerdle B. Factors associated with chronic pain and fat mass in a population-based cohort. Pain Rep. 2018;3(5):e676. doi:10.1097/PR9.0000000000000676

23. Walsh TP, Arnold JB, Evans AM, Yaxley A, Damarell RA, Shanahan EM. The association between body fat and musculoskeletal pain: a systematic review and meta-analysis. BMC Musculoskelet Disord. 2018;19(1):233. doi:10.1186/s12891-018-2137-0

24. Dunn LK, Yerra S, Fang S, et al. Preoperative opioid use is associated with increased length of stay and persistent opioid use following spine surgery. J Spine Surg. 2018;4(2):167–174. doi:10.21037/jss.2018.05.15

25. Line KM, Terkawi AS, Tsang S, et al. Preoperative opioid use as a predictor of spinal cord stimulation outcomes. Neuromodulation. 2021;24(3):508–516. doi:10.1111/ner.13267

26. Deyo RA, Hallvik SE, Hildebran C, et al. Association between initial opioid prescribing patterns and subsequent long-term use among opioid-naive patients: a statewide retrospective cohort study. J Gen Intern Med. 2018;33(7):1226–1234. doi:10.1007/s11606-017-4201-8

27. Grace PM, Strand KA, Maier SF, Watkins LR. Suppression of voluntary wheel running in rats is dependent on the site of inflammation: evidence for voluntary running as a measure of hind paw-evoked pain. J Pain. 2014;15(1):121–128. doi:10.1016/j.jpain.2014.01.001

28. Hernandez I, He M, Brooks MM, Saba S, Gellad WF. Concurrent opioid-benzodiazepine prescribing and adverse outcomes. Pain Med. 2018;19(11):2201–2209. doi:10.1093/pm/pnx283

29. Olopoenia A, Onukwugha E, Simoni-Wastila L, et al. Opioid-benzodiazepine co-prescription and persistent opioid use. Pain Med. 2022;23(3):504–513. doi:10.1093/pm/pnab238

30. Jung YH, Kim H, Jeon SY, et al. Polypharmacy and outcomes after interventional pain procedures: a multicenter cohort study. Pain Pract. 2024;24(1):134–143. doi:10.1111/papr.13306

31. Yi MH, Wu YJ. TLR4-dependent microglial activation mediates chronic pain maintenance: optogenetic evidence. Nat Neurosci. 2021;24(6):832–842. doi:10.1038/s41593-021-00852-0

32. Lotsch J, Ultsch A. Machine learning in pain research. Pain. 2018;159(4):623–630. doi:10.1097/j.pain.0000000000001118

33. Neblett R, Cohen H, Choi Y, et al. The Central Sensitization Inventory (CSI): establishing clinically significant values for identifying central sensitivity syndromes in an outpatient chronic pain sample. J Pain. 2013;14(5):438–445.

